# Machine Learning Based Clinical Decision Support System for Early COVID-19 Mortality Prediction

**DOI:** 10.1101/2020.08.19.20177477

**Authors:** Akshaya Karthikeyan, Akshit Garg, P. K. Vinod, U. Deva Priyakumar

## Abstract

The coronavirus disease 2019 (COVID-19) is an acute respiratory disease that has been classified as a pandemic by World Health Organization (WHO). The sudden spike in the number of infections and high mortality rates have put immense pressure on the public medical systems. Hence, it’s crucial to identify the key factors of mortality that yield high accuracy and consistency to optimize patient treatment strategy. This study uses machine learning methods to identify a powerful combination of five features that help predict mortality with 96% accuracy: neutrophils, lymphocytes, lactate dehydrogenase (LDH), high-sensitivity C-reactive protein (hs-CRP) and age. Various machine learning algorithms have been compared to achieve a consistent high accuracy across the days that span the disease. Robust testing with three cases confirm the strong predictive performance of the proposed model. The model predicts with an accuracy of 90% as early as 16 days before the outcome. This study would help accelerate the decision making process in healthcare systems for focused medical treatments early and accurately.

## Introduction

The outbreak of the novel coronavirus disease 2019 (COVID-19) caused by the virus SARS-CoV-2 began in Wuhan, China in December 2019. Since then, it has rapidly spread around the world. As of November 4, 2020, WHO reported a total of about 47 million confirmed cases and more than one million deaths worldwide. At this stage of the pandemic, most estimates of fatality ratios have been based on cases detected through surveillance and calculated using crude methods, giving rise to widely variable estimates of case fatality rate by country. The sudden rise in cases has put immense pressure on the healthcare systems due to limited resources. Identification of the key bio-markers of mortality is crucial because it helps in understanding the relative risk of death among patients and therefore guides policy decisions regarding scarce medical resource allocation during the ongoing COVID-19 pandemic.

Symptoms of COVID-19 are very similar to the common flu that include fever, cough and nasal congestion(1). As the pandemic spread, other symptoms such as loss of taste and smell (anosmia) have emerged(2). Severe cases can lead to serious respiratory disease and pneumonia. Those most at risk are the elderly and people with underlying medical issues/ comorbidities, such as cardiovascular diseases and diabetes (3, 4, 5). As the disease spreads around the world, more symptoms and features that affect patient mortality are being realized. Having such a large set of features that are affected by the disease makes it hard to understand which ones have a greater impact on patient mortality. Machine learning methods can aid by analyzing large sets of data quickly and providing models that assess risk factors accurately.

Artificial intelligence aids the decision making process by giving predictions that are fast, accurate and interpretable. Several studies have used machine learning algorithms for COVID-19 prognosis which can be utilised to do better resource planning. (6, 7, 8, 9, 10). Wang et al.(11) proposed two different models based on Clinical and Laboratory features to predict mortality of COVID-19 patients. The Clinical model developed with Age, history of hypertension and coronary heart disease showed AUC of 0.83 (95% CI, 0.68-0.93) on the validation cohort. The laboratory model developed with age, high-sensitivity C-reactive protein (hs-CRP), peripheral capillary oxygen saturation (SpO2), neutrophil and lymphocyte count, D-dimer, aspartate aminotransferase (AST) and glomerular filtration rate (GFR) had better discriminatory power with AUC of 0.88 (95% CI, 0.75-0.96) on the validation cohort. The Validation cohort consisted of 44 COVID-19 patients of which 14 died and 30 survived. XGBoost and backward step-down selection were used for feature selection followed by a multivariate logistic regression for the classification. The clinical model can prove to be useful given the ease of data collection of all the three features. Shang et al.(12) established a scoring system of COVID-19 (CSS) to split patients into low-risk and high-risk groups. Here, high-risk group patients would have significantly higher chances of death than those of the low-risk group. Multivariable analysis and coefficients of lasso binary logistic regression were used to do feature analysis and to establish a prediction model. Eight different variables including age and blood parameters were used to generate a model which showed good discriminative power with an AUC of 0.938 (95% Cl, 0.902-0.973) on the independent validation cohort. Xie et al.(13) identified SpO2, Lymphocyte Count, Age and Lactate dehydrogenase (LDH) as a set of important features to generate a mortality prediction model. They used multivariable logistic regression for the classification task which gave an AUC of 0.98 on the independent validation set. Using this they established a nomogram to generate probability of mortality. An interpretable mortality prediction model for COVID-19 patients has been proposed by Yan et al.(14) where they analyzed blood samples of 485 patients from Wuhan, China, and proposed an XGBoost based solution for mortality prediction. The proposed clinically operable single tree XGBoost model used three crucial features-LDH, lymphocytes and hs-CRP. The decision rules with the three features along with their threshold were devised recursively. This provided a interpretable machine learning solution along with an accuracy of atleast 90% for all days.

The papers published have not presented the consistency of their results for different days to outcome. For example, Yan et al. analysed their model performance and showed 94% accuracy at 7 days before day of outcome. However, it’s possible that their results are biased towards the high number of samples near to the day of outcome. A model should be able to predict mortality independent of days to outcome consistently because a patient could be in any stage during the span of the disease. It is also desirable to be able to do risk stratification during the early days of infection, which is helpful in devising treatment strategies appropriately.

In this study, we have used the dataset provided by Yan et al. for developing our models. We created a solution using XGBoost feature importance and neural net classification. A strong combination of five features was chosen as the key biomarkers for prediction. The neural net provides a high predictive performance (96% accuracy), while the XGBoost feature importance graph adds interpretability to the model. Analysis of the features’ graphs showing trends in progression provide additional insights into the features as well. Various algorithms and robust testing were implemented to establish a strong confidence in the model. Our model proves to be extremely accurate and consistent through the days spanning a patient’s disease. This would help in faster diagnosis with fewer number of features and higher confidence. Healthcare systems can use the model to optimize treatment strategy by focused utilization of resources.

## Results

### Machine Learning Pipeline

Figure 1 depicts the overall pipeline used in this study for performing the mortality prediction task. As mentioned above, we have used the dataset provided by Yan et al for developing the machine learning models.(14) All the models have been trained on samples from all days from this dataset for mortality prediction irrespective of the number of days to outcome. Following data preprocessing, XGBoost classifier was used to obtain feature importance, and a neural network was used for feature selection. The optimum combination of features thus obtained was then used to train various supervised machine learning classification models. The trained models were then tested based on three different ways, with each having its own strengths. Five-fold cross validation was utilized to assess the predictive ability and statistical significance of the models. Assessment of the developed models was done based on different metrics whose mean and standard deviation are reported below. Further detailed account of the step-by-step procedure are presented in the Methodology section later.

**Figure 1.**
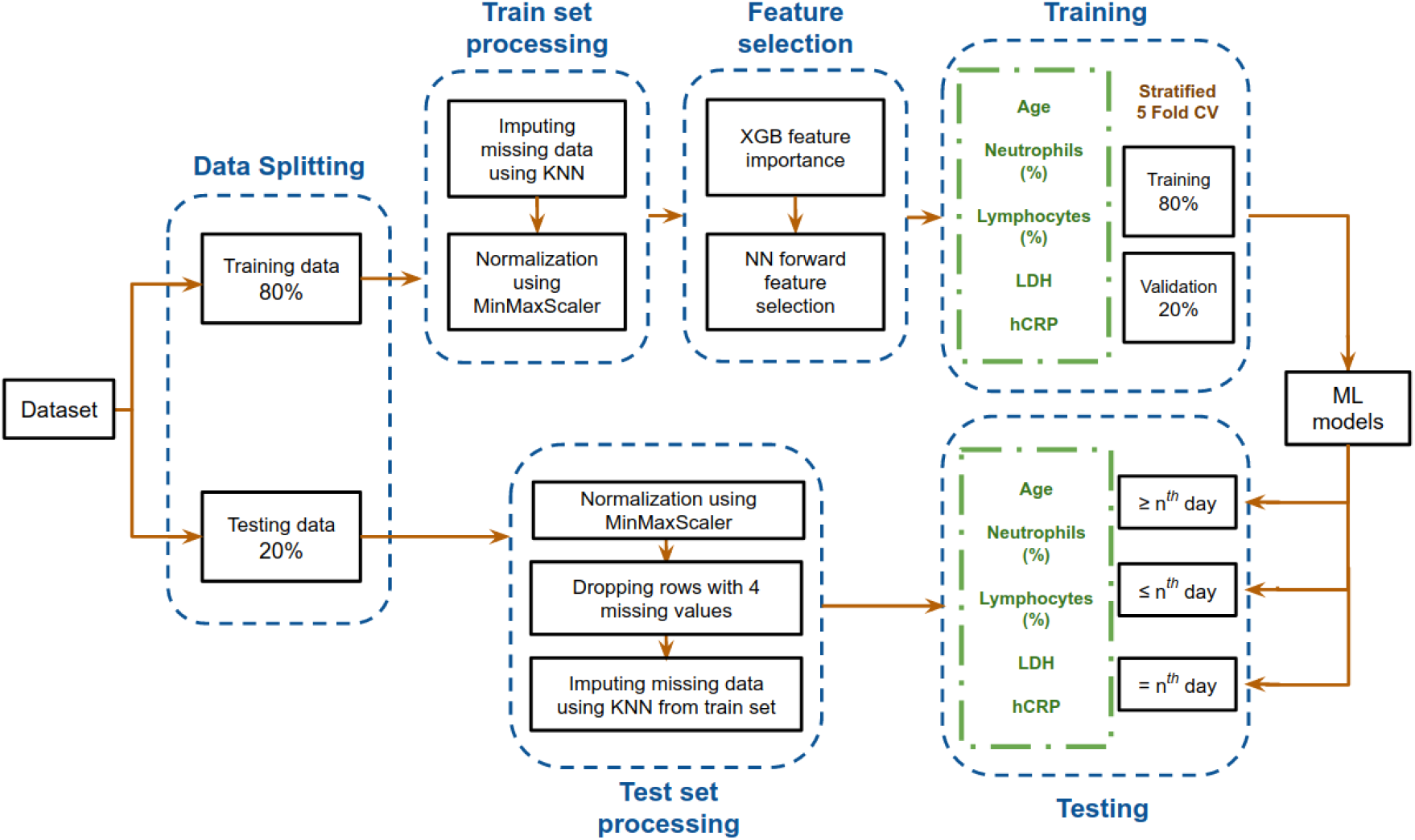
Flowchart depicting the model development pipeline used in this study.

### Data Splitting for Unique Patient Segregation

The dataset after pre-processing consists of 1766 datapoints corresponding to 370 patients, out of whom 54.3% recovered and 45.7% succumbed to COVID19. The number of datapoints for each of the patients range from 1 to 12 that were collected during different days before one of the two outcomes. To ensure exclusivity of patients in the training and testing sets, 80% of the patients were randomly chosen for the training set and remaining 20% of the patients were chosen for the testing set. Unique patient segregation is important since including datapoints from a single patient in the training and test sets may lead to bias. Such a unique 80:20 split gave us a training set comprising of 1418 data points and testing set comprising of 348 data points. The class distribution across the days to outcome is shown in Figure 2. The classes are spread across all the days in good ratios and are comparable. Since our aim is to develop mortality prediction model that is independent on the days to outcome, all the readings of the patients are considered as unique data-points for further examination.

**Figure 2.**
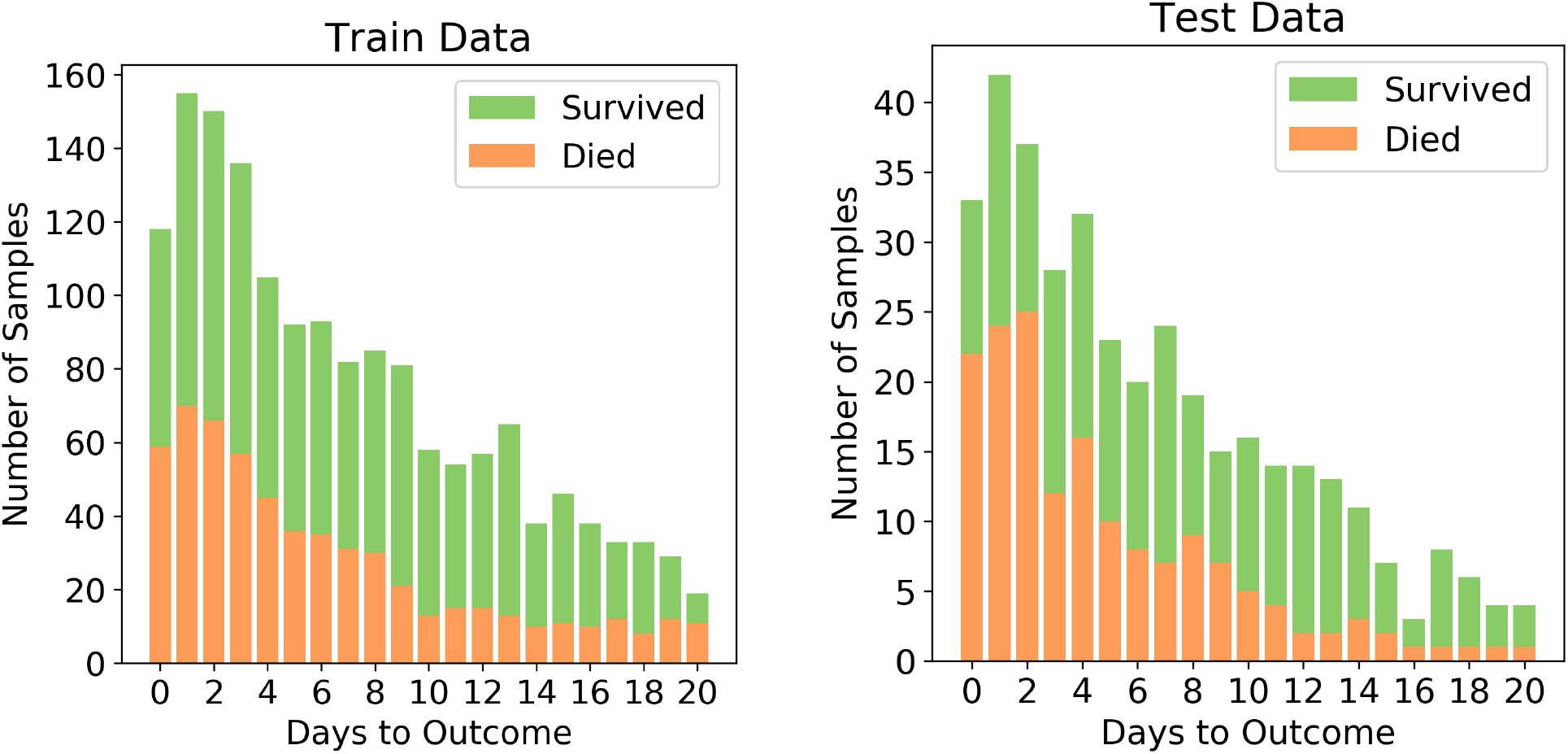
Distribution of the two classes in the train and test sets after splitting

In this study, results with and without KNN imputation are compared. KNN algorithm is useful for matching a data-point with its closest *k* neighbours in a multi-dimensional space. <u>*Imputed test set:*</u> Each data-point in the test set was imputed using the values corresponding to the selected five features from their ten nearest neighbours in the train set. The resulting test set contained 213 samples belonging to 71 patients. The dead/total patient ratio of 0.563 shows that there is a good representation of both classes in the test set. This ensures that the results are reliable for both classes. <u>*Non-imputed test set:*</u> To assess the performance of the algorithms when the test set has no synthesized values, we dropped the rows that had missing values in any one of the five features. The resulting test set contained 115 samples belonging to 65 patients with a dead/total patient ratio of 0.513.

### Identification of Key Biomarkers

Given, especially, the severity and rapid spike in COVID-19 infections and resulting fatalities, a large number of lab-tests is required to assess patients’ medical conditions. Feature selection by which the most important and crucial biomarkers that aid in risk assessment are identified, is an important exercise because acquiring fewer lab tests means faster and efficient decision making processes.

#### XGBoost feature importance

The relative importance of available features were determined using the gradient boosting algorithm, XGBoost. It was observed that the top four features are neutrophils (%), lymphocyte (%), LDH and hs-CRP in the given order. Figure S1 shows the order of relative importances of all the features originally considered. The list of features was sorted in the descending order of importance. ‘Age’ was then added to the top of the feature importance list owing to its extreme ease of data collection and studies(3, 4, 5) showing that its an important factor in determining disease progression of COVID-19 patients.

#### NN feature selection

The AUC scores of each set of features during forward selection using a neural network was plotted. The aim of this exercise was to maximize AUC score and minimize the number of features selected for model development. It was observed that the mean AUC score at five features was 0.95. Adding the sixth feature did not increase the AUC significantly. Observed AUC with respect to the number of features selected for modeling is given in Figure S2. It was also noticed that dropping ‘age’ feature hampered the performance of the model. Hence, the features selected for this study are age, neutrophils (%), hs-CRP, lymphocyte (%) and LDH.

### Predictive Ability of the Model

Six different algorithms, namely neural net, SVM, logistic regression, random forests, XGBoost and decision trees, were used to develop the classification models. Initially, each algorithm after stratified five-fold cross validation was tested on the test set, which included samples from all days. Figure 3 shows the accuracy, F1 Score and AUC of all the developed models using the six different methods. Our aim is to choose the most accurate model that is highly capable of distinguishing the two classes. It was observed that the best model is the neural net as it performed better than rest in terms of accuracy (96.53% *±* 0.64%) and F1 score (0.969 *±* 0.006). It has a high AUC score (0.989 *±* 0.006), showing good distinguishing capacity for the features of both classes. Hence, the results and discussions presented in the rest of the manuscript will be based on those obtained using the neural network unless otherwise mentioned.

**Figure 3.**
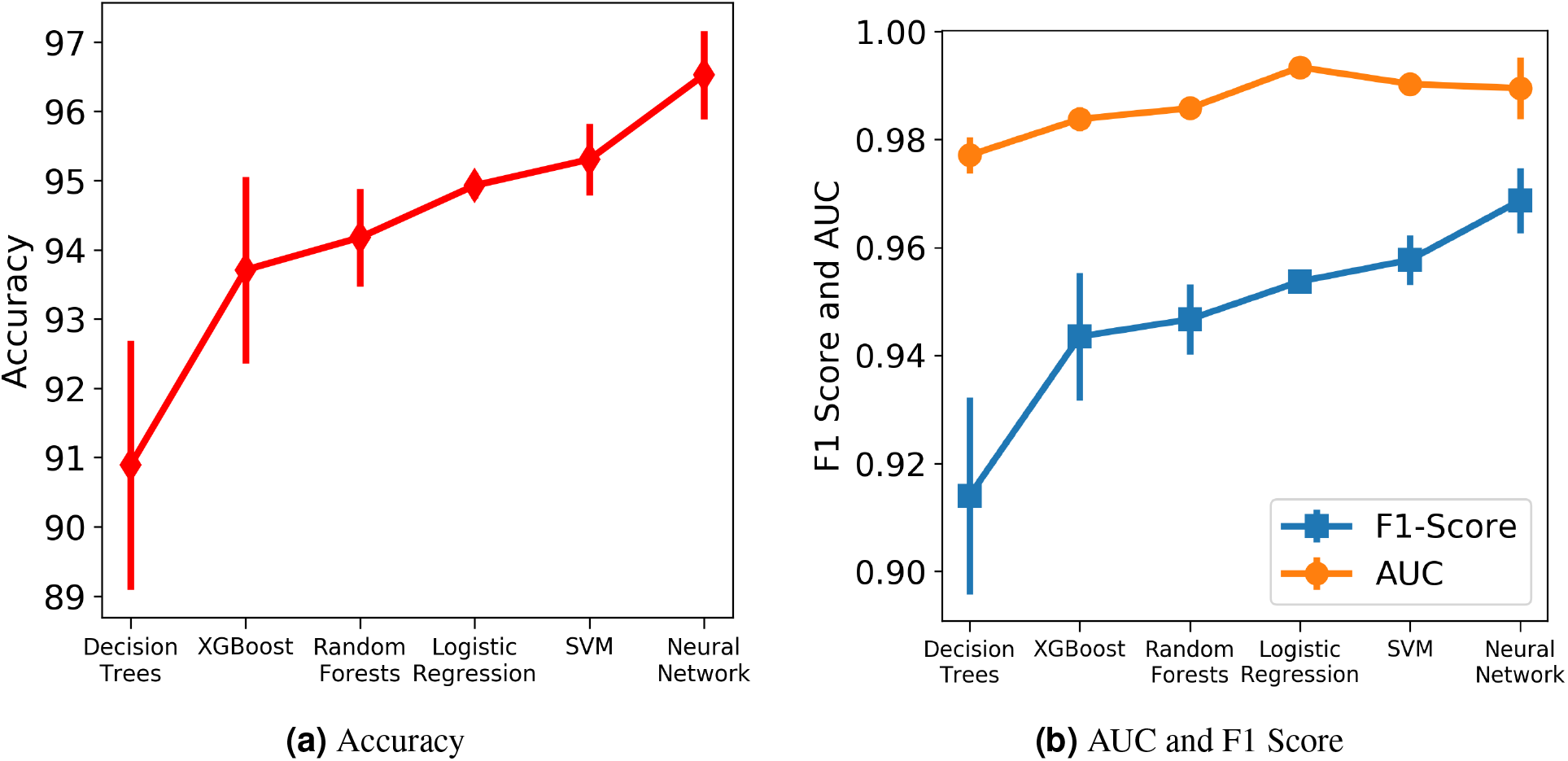
Comparison of the performance of different machine learning algorithms assessed using different metrics. The vertical lines denote the standard deviations.

The robustness and applicability of the model at different settings were further tested by considering three different cases (Cases 1, 2 and 3). The first case investigates the performance of the model when the test set consists of data only if *n* or less number of days are left till the day of outcome. (Figure 4a) Testing was done with respect to different values of *n*. From Figure 4b, it was observed that the accuracy is consistently high for *n* up to 17 days if only *n* or less days are left till the outcome. For the value of range of *n* between 0 and 17 days, the accuracy values were in a close range of 97.1 to 99.0% indicating the high predictive nature of the model. The number of samples after day 17 is relatively very few to affect this accuracy at later days, hence all the analysis have been done up to only 17 days. From Figure 4c, we observe high and consistent AUC scores (best = 1 and worst = 0.99) and F1 scores (best = 0.99 and worst = 0.97) across various days. This shows that the model performs consistently with an accuracy of at least 97% if any number of days are left till the outcome.

**Figure 4.**
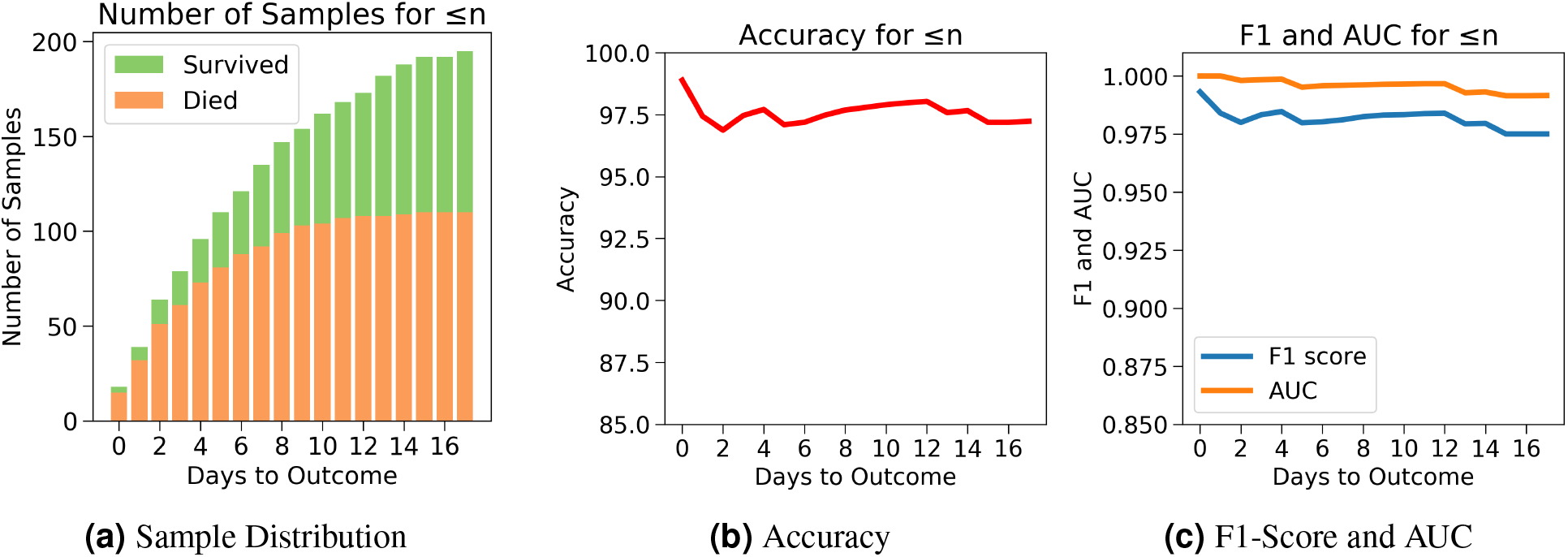
Case 1: Number of days to outcome less than or equal to *n*. (a) The class-wise distribution of the cumulated data-points (≤ *n*^*th*^*day*) for all samples in the imputed test set. (b) Accuracy of the model evaluated for different days to outcome. (c) F1-score and AUC of the model evaluated for different days to outcome.

The drawback of Case 1 is that it is possible for the samples nearer to the outcome to dominate the results since there are more samples nearer to the outcome. Case 2, which examines the performance of the model for *n* or more number of days in advance, was considered to address this drawback. One does not know when the day of outcome is going to be; hence it is important to analyze the model’s performance with respect to any day before the day of outcome. This would also help to assess the performance of the model over the span of the disease. Since the number of samples decreases with respect to the number of days before outcome, every cumulation can only be dominated by the samples of the day closest to the outcome. (Figure 5a) Hence, this gives a more accurate overview. Figure 5b agrees with our intuition, that it gets harder to predict the outcome as we go farther from the day of outcome. Nevertheless, the model starts with a high accuracy of 96.5% at the day of outcome and stays quite consistent. The lowest accuracy of 88% was observed when the model predicts 15 or more days in advance. From Figure 5c a similar consistency was observed with the AUC and F1 scores. The AUC starts with 0.99 at the day of outcome and reaches its lowest point on day 15 with 0.96. F1 scores start with 0.97 and reach its lowest point 0.84 on day 13. However the model predicts with a high accuracy of 95.7% when *n* ≥1 week indicating strong performance.

**Figure 5.**
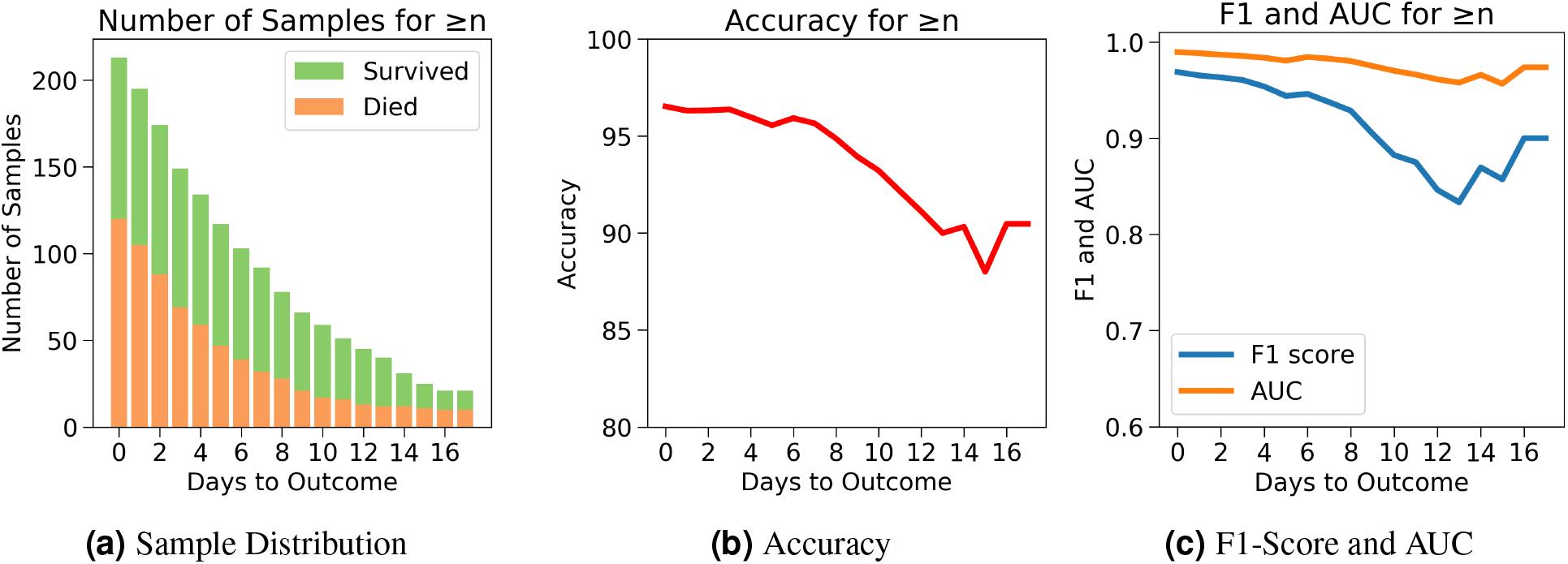
Case 2: Number of days to outcome greater than or equal to *n*. (a) The class-wise distribution of the cumulated data-points (≤ *n*^*th*^*day*) for all samples in the imputed test set. (b) Accuracy of the model evaluated for different days to outcome. (c) F1-score and AUC of the model evaluated for different days to outcome.

Next, for even more rigorous testing, we introduce Case 3 which assesses the performance of the model on exactly the *n*^*th*^ day before the day of outcome (Figure 6a). Here, no sample from any other day can influence the results. Even though the accuracy fluctuates through the days, Figure 6b shows that the model predicts the outcome with an accuracy of 98.9% on the day of outcome and reaches its lowest value of 92.85% on day 5. The AUC and F1 scores follow a similar trend by starting at 1 and 0.99 respectively on day of outcome and reaching its lowest score on day 5 with 0.95 and 0.94 respectively (Figure 6c). It’s possible that the 100% accuracy for day 7-12 could be due to the less number of samples, but its worth noting that both classes were present and the model consistently predicted them all correctly for day 7-12 with AUC and F1 scores at 1. Figures S3-S7 show the performance of the other five algorithms after testing with the three cases.

**Figure 6.**
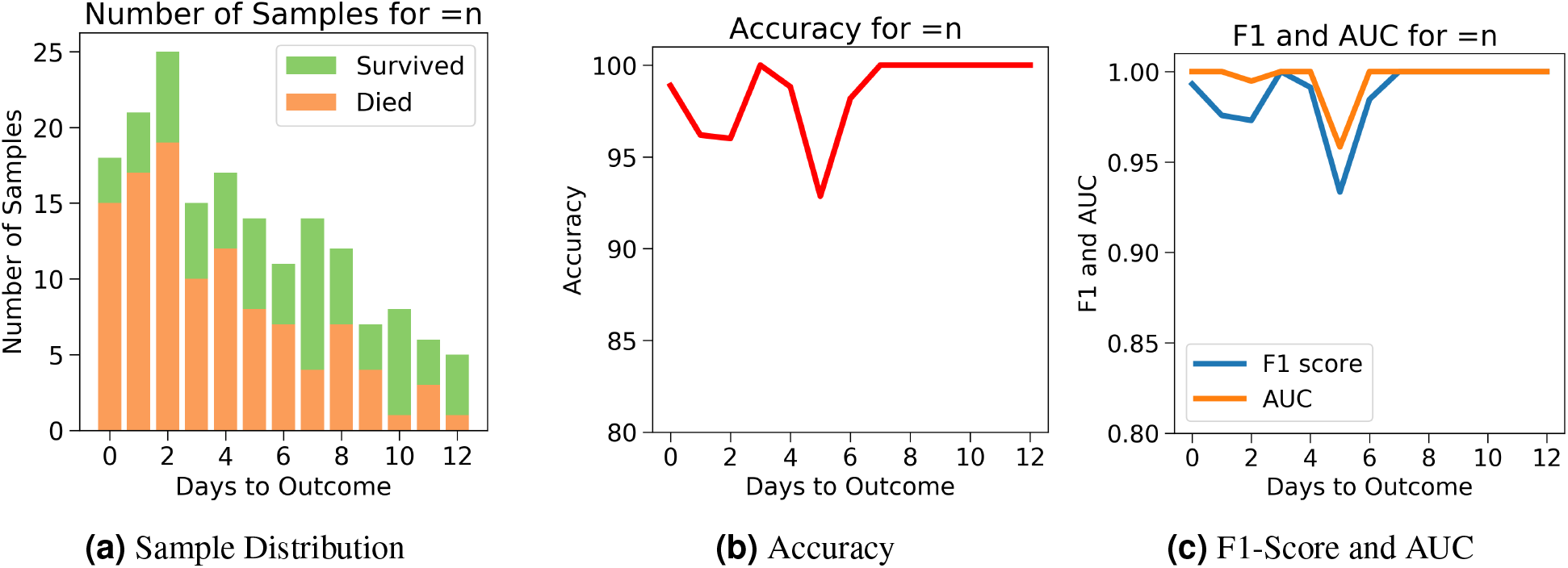
Case 3: Number of days to outcome equal to *n*. (a) The class-wise distribution of the cumulated data-points (≤ *n*^*th*^*day*) for all samples in the imputed test set. (b) Accuracy of the model evaluated for different days to outcome. (c) F1-score and AUC of the model evaluated for different days to outcome.

As mentioned above, the models and their performances presented above were trained based on the test data that was imputed using the training set. Models were also trained using the six different algorithms using the test data without any synthetic data (without imputation) by just simply dropping the data points with missing data. From Table S2, it can be observed that again the neural network performed well on the test set with an accuracy of 94.61% *±* 0.85%, AUC as 0.98 *±* 0.01 and F1 Score of 0.95 *±* 0.01. Logistic Regression is the next best performing algorithm with results close to neural network. For robust testing, we again used the three cases (Cases 1, 2 and 3) as discussed above to determine the predictive performances. Figures S8-S15 show the performance of all the models after testing with the three cases.

## Discussion

The COVID-19 pandemic has put an immense pressure on the healthcare systems around the globe due to the rapid rise in the number of infections. In these times, it is extremely crucial to assess risk such that critical resources can be mobilized to treat patients progressing to severe stages. Focused medical treatments can be administered only when there is a clear understanding of the risk factors that influence the mortality the most. Due to the recent rise of the disease, new features that affect the progression of the disease are continuously being inquired. Machine learning methods are capable of discerning useful patterns in large dimensional data. This study reports machine learning model that is expected to aid in the decision making process of identifying patients who are at high risk with high accuracy.

In this study, XGBoost feature importance and neural network were utilized to find the right balance between high AUC score and low number of features selected for developing the ML models. In this process, five features were chosen to create a powerful combination for mortality prediction. The selected five features include neutrophils (%), hs-CRP, age, lymphocyte (%) and LDH. Each of these features have been identified as predictors of mortality associated with the COVID-19 disease. (9, 11, 12, 13, 14, 15, 16, 17) Age has been identified as an important factor in COVID-19 disease progression and hence it has been included in all the models here.(3, 4, 5) Patients aged ≥60 years had a higher rate of respiratory failure and needed more prolonged treatment than those aged <60 years,(3) implying that the elderly showed poorer response to treatments than the younger age group. Older patients (age ≥80 years) had a risk of 41.3% of having severe or critical condition upon contracting COVID-19 while younger patients (age <20 years) had a lower risk of only 4.1%.(4) Older people are also more susceptible to co-morbidities which has been identified as another independent risk factor for COVID-19 disease prognosis.(5)

Neutrophils are a type of white blood cells and are the first line of defence in the inflammatory response.(18) Elevated levels of neutrophils (neutrophilia) suggest that the body is infected. Figure 7b shows that patients who died had significantly higher levels of neutrophils (%) compared to patients who survived. Lymphocyte consist of commonly known B cells, T cells and NK cells. Figure 7c shows that people who died had significantly lower levels of lymphocyte (%) than those who survived. Neutrophil to Lymphocyte Ratio (NLR) has been identified as an independent risk factor for critical illness in COVID-19 patients.(19)

**Figure 7.**
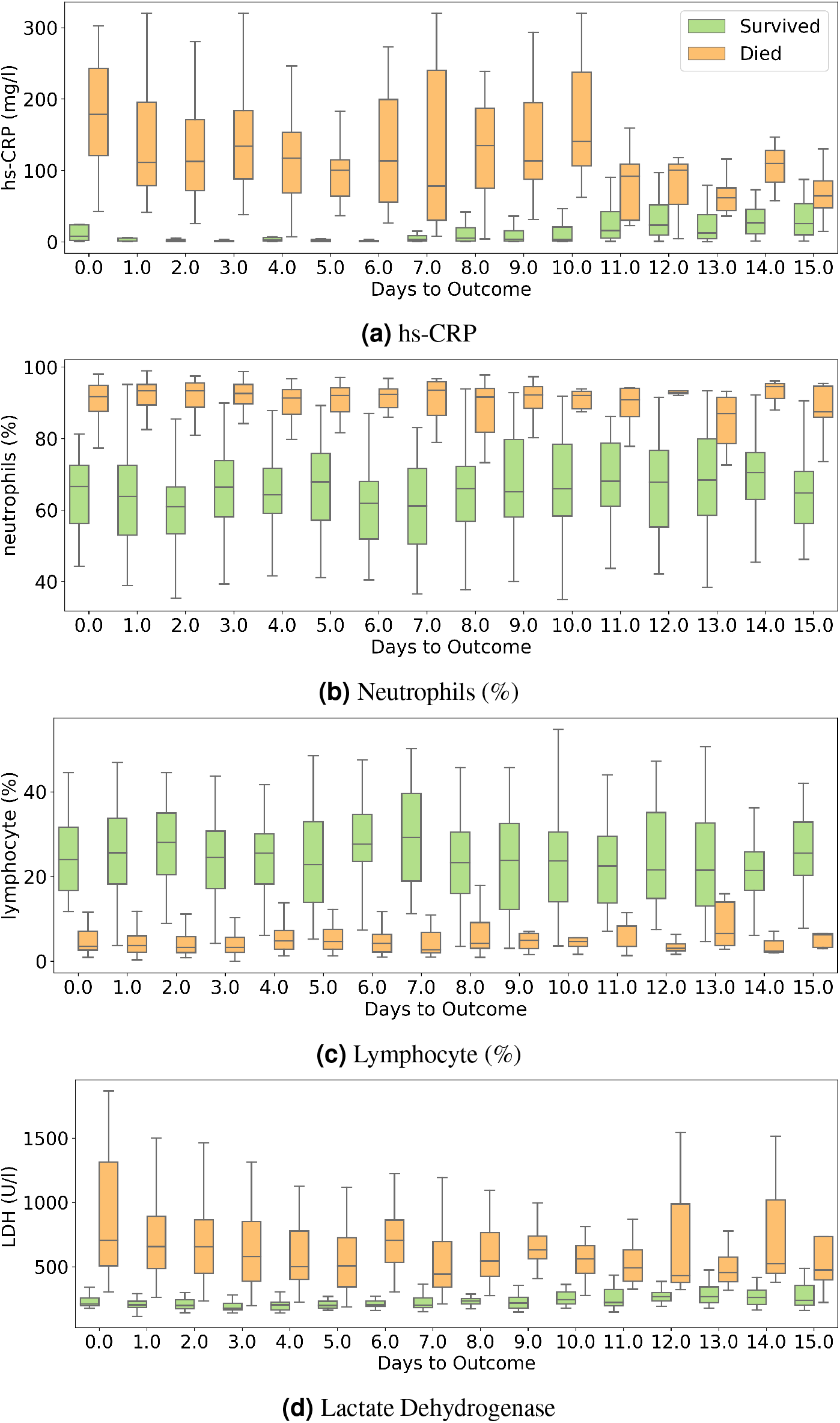
Box and whisker plot showing the variations of four selected features with respect to the days to outcome.

Figure 7d shows that patients who died had higher LDH levels than those who survived. High LDH levels are also linked to co-infection and possibly predict prognosis in severe bacterial infections.(20) LDH has been identified as an important indicator of lung damage or inflammation.(21) Several studies have identified LDH as an important factor to study COVID-19 disease progression and have linked high levels of LDH to higher risk or severity and fatality.(22, 23, 24) While it is helpful to provide cutoff values of the features identified here for direct applicability(14), this can vary depending on the type of methods used for measurement. For example, LDH levels cut-off can vary depending on measurement techniques.(25) Yan et al.(14) clarified that Tongji Hospital used the conversion of lactate to pyruvate (L → P) with concomitant reduction of NAD^+^ to NADH.(26) According to the guidelines of the kit used, the normal range is ≤ 250 U/l^−1^ in adults. Also, blood samples with haemolysis were not added in the dataset.(26)

hs-CRP is produced in the liver that responds to a wide range of health conditions, leading to inflammation. Figure 7a shows that people who died had higher levels of hs-CRP than those who survived. Studies have found that higher hs-CRP levels correlates with lower pulmonary functions (27, 28) and patients with Chronic Obstructive Pulmonary Disease (COPD) have been found to be with higher hs-CRP level compared to normal population.(29, 30) hs-CRP has also been identified as an important factor to facilitate triage of COVID-19 patients(31). Very high hs-CRP level show the development of severe bacterial superinfection, which is expected to be a frequent complication in critical patients with COVID-19, and potentially a reason for increased mortality. Identifying patients at higher risk of superinfections or other complications before observing substantial increases in hs-CRP and LDH levels may help in treating them more efficiently.(20)

We compared several machine learning models for their predictive performance with the selected 5 features. The test set had a good representation of both the survived (43.66%) and dead (56.34%) patients. To realize the actual predictive performance of the models and understand how confident we can be with the results, robust testing was carried out using 3 cases. It was observed that the neural net consistently performed better than the rest of the models. Neural network was able to predict patients’ mortality with an accuracy of 96.53% and an F1-Score of 0.969 on a test set of 213 samples spread across multiple days during the span of the disease. The highly accurate and consistent performance of the neural net based model after robust testing with the five chosen features gives a strong confidence on the model.

Other machine learning models involving trees and regression algorithms performed with an average accuracy of 94%. This shows that the five selected features are extremely influential on patient mortality. Figures 7 clearly show how the selected features show a difference in trend for the two classes though there is a significant overlap between the two sets of data. This explains the decent performance of simple algorithms like Logistic Regression and SVM. Neural Networks move a step ahead by distinguishing the features for the two classes to a greater accuracy and greater consistency through the days. Although the neural networks take time to train, once trained they are able to produce the results at speeds comparable to simple algorithms.

We have tested using stratified five-fold cross validation and ensured a good representation of both the classes in the test set.(32) Yan et al.(14) only tested their model using Case 1 whereas our model was tested using two more practical cases, and performed more accurately and consistently on Case 1 as well. The proposed models have been developed based on the data from patients exclusively from a single hospital from Wuhan forcing it to have certain biases including towards the viral strain found in Wuhan. Mutations might have changed the disease progression patterns in other populations.(33, 34, 35, 36, 37) Nevertheless, studies done on other cohorts have also identified these features as key predictors.(15, 16, 17) After experimenting with various algorithms, we observed that there is a trade-off between accuracy and interpretability. The best performing model, neural network, works like a black-box. However, XGBoost importance graph adds interpretability to it. If the model could be improved using data from diverse sources, implementing either of the types of models in the clinical setting is possible. The currently proposed models are accurate enough to **(d)** Lactate Dehydrogenase capture the mortality rates and hence can help in targeted caring of high risk patients.

Features other than the five used for developing the reported models were further analyzed to identify the ones that show different trends with respect to the outcome during the span of the disease. Figures (a)-(i) in figure S16 show the progression of nine other features that seem to show trends with respect to number of days to outcome. Calcium and serum potassium could be used to predict days until death for critical patients, as it shows an increasing trend in value towards the day of outcome. Platelet count can be used to analyze trends of features in patients belonging to both the classes. The dataset used for this work is insufficient to conduct such a study due to its size and density. We believe that with a larger dataset it may be possible to analyze these trends more meaningfully and possibly predict the number of days to outcome. This can potentially scale up the resource planning by many folds and could prove to be of great significance.

## Summary

In summary, this study reports the identification of powerful combination of five features (neutrophils (%), hs-CRP, age, lymphocyte (%) and LDH) that helps in accurately assessing the mortaility rates of COVID-19 affected patients. Different machine learning models have been developed to compare and predict mortality most accurately. The neural network predicts the survival rates with 96% accuracy for all days during the span of the disease and with 90% accuracy for more than 16 days in advance. XGBoost feature importance provides interpretability to the model that may be relevant in the clinical setting. The robustness of the proposed model was thoroughly tested with three different scenarios. The performance metrics obtained instill great confidence on the proposed model. Other possible features that have predictive capability are identified, however will need data from diverse sources to further confirm their relevance and to possibly improve the model.

## Methodology

### Dataset and Preprocessing

All data for feature analysis, training and testing were taken from Yan et al.(14) This dataset includes 2,779 electronic records of validated or suspected COVID-19 patients from Tongji Hospital in Wuhan, China. The initial dataset comprised of the time series data of 375 COVID-19 patients with 74 biomarkers along with data sample time, admission time, discharge time and outcome (survived or dead). Yan et al. used only data of the final samples of each patient for training and testing.(14) In our study, we have considered the samples from all the days of each patient for training and testing. For each patient, there were multiple rows representing readings taken on different days. Some days also had multiple readings taken at different times. All such rows representing same day readings of a patient were combined together to create a single data point for each unique day of the patient. In cases where there existed features with multiple recordings taken in a single day at different times, the readings which was taken the earliest in that particular day is considered for generating the combined single data point. This is because we require the model to learn the features that predict mortality at the earliest. Features which had missing values in more than 70% of the instances were dropped and were not used for further analysis. A new column ‘Number of days till Outcome’ is added to signify how many days are left for a sample to reach the day of outcome. This is calculated by subtracting the day of reading from the day of discharge/death. After data generation and processing, our dataset had 201 patients who survived and 169 patients who died. The features were analyzed with respect to the two classes and selected graphs are provided in Figure S17.

Missing values in the training set were imputed using K-Nearest Neighbour algorithm. The value for imputation was calculated by averaging out the values of 10 nearest neighbours weighted with respect to inverse of euclidean distance. For normalising the training data Min-Max scaling was used. Min-Max scaling was used since most features don’t fit a normal distribution and to ensure that the structure of the data-points in each feature remained intact.

### Evaluation Metrics

The predictive performance of the supervised models was assessed using the following metrics. Here, TP, TN, FP, and FN stand for true positive, true negative, false positive and false negative rates respectively).

#### AUC (Area under ROC curve)

AUC is the area under receiver operating characteristic curve (ROC curve in short). AUC provides an aggregate measure of performance across all possible classification thresholds. The ROC curve plots two parameters:

- True Positive Rate

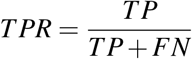
- False Positive Rate

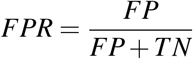

#### Accuracy

Accuracy is an important metric for classification models. In this study, the test dataset is not unbalanced, hence this will give a good idea about the model’s predictive performance.

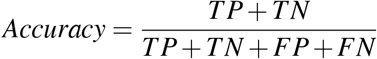

#### F1 Score

F1 score is the harmonic mean of precision and recall.

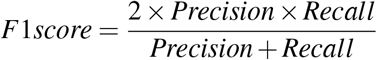

where,

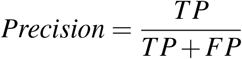

and,

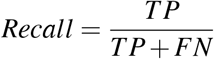

### XGBoost feature importance

To evaluate the biomarkers that have the most influence on the outcome, XGBoost was used to get the relative importances. XGBoost is a powerful machine learning algorithm that estimates features that are the most discriminative of model outcomes. The accumulated role of each individual feature in each decision step in trees is used to calculate its final importance. The average importance of the features were found through 100 iterations of randomly selecting 80% of the samples in the training set. The parameter settings for XGBoost of maximum depth was set to 3, learning rate was set to 0.2 and regularization parameter *α* was set to 5 with logistic regression as the objective. All the other parameters carry their default values. ‘Age’ was then added to the top of the feature importance list due extreme ease of data collection.

### NN feature selection

After determining the order of importance, neural network comprising of two hidden layers was used to find the optimal number of features required for mortality prediction. Neural networks can learn complex relations between the features. Theoretically, shallow neural network is capable of performing as good as or better than logistic regression models. The average AUC of each set during feature selection was calculated using stratified 5 fold cross validation. The performance of each set of features was then compared to select the optimum one.

### Training

To compare the performances of various machine learning algorithms, we tried six different models as listed below. The most optimum feature set was used as inputs to these models. GridCV with Stratified 5-Fold cross validation was used for extensive hyper-parameter tuning of Logistic Regression, Random Forests, XGBoost, Support Vector Machine and Decision Trees.

1. <u>Neural Network:</u> The Neural Network trained for feature selection had two hidden layers with ReLU activation. Binary Cross Entropy with Logits was the loss function. Adam optimizer with learning rate 0.001 and Reduce On Plateau scheduler with patience as 5 was utilized to update the weights and learning rate. Figure 8 shows the architecture of the neural network. A similar Neural Network with few differences was used for predicting mortality-learning rate was set as 0.00001 and patience of the scheduler was set as 50 to ensure good fitting on the training data. The cutoff for classification with was evaluated with respect to the validation set and F1-Score was used as the metric for performance comparison. In case multiple cutoffs gave the same F1-Score, the cutoff closest to 0.5 was chosen.

**Figure 8.**
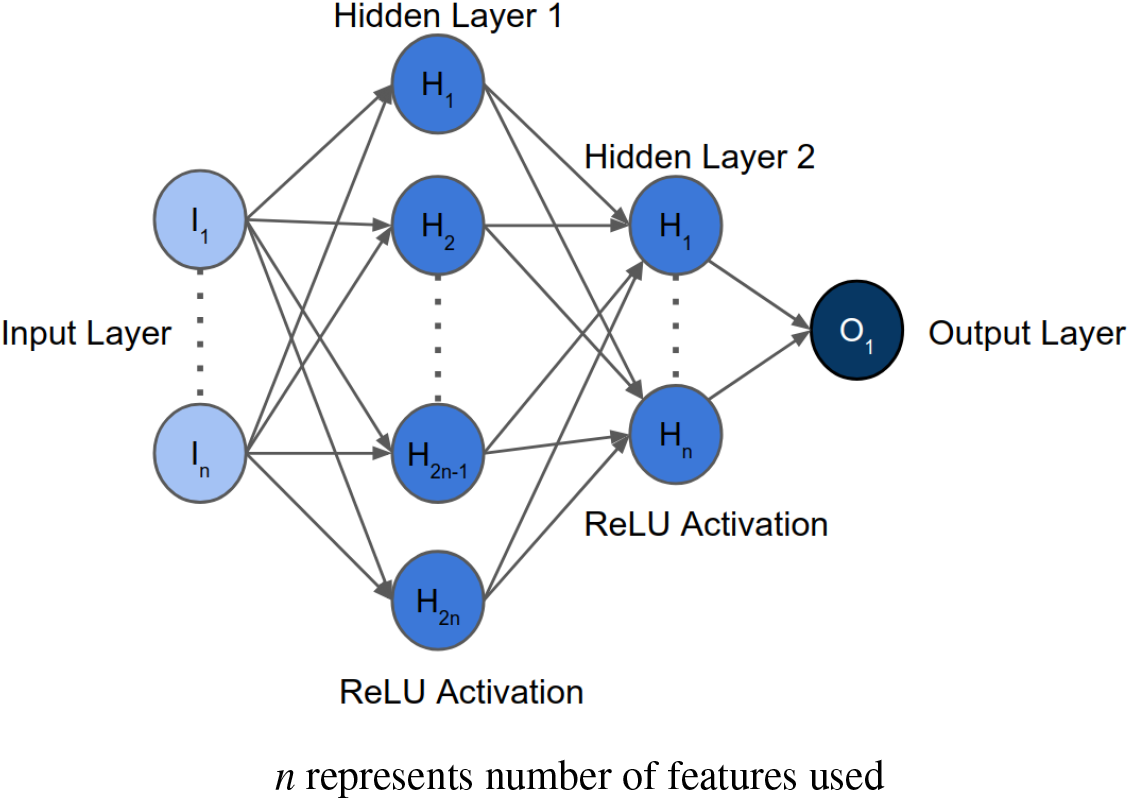
Architecture of the neural network implemented for feature selection
2. <u>Logistic Regression:</u> Logistic Regression was trained with ‘liblinear’ solver due to small dataset size, L1 penalty, tolerance for stopping criteria set as 0.0001, inverse of regularization strength C set as 10 and intercept scaling set as 1.
3. <u>Random Forests:</u> Random Forests was trained with gini criterion, maximum depth set as 9, minimum number of samples at a leaf node set as 1 and number of trees set as 90.
4. <u>XGBoost:</u> XGBoost was trained with objective set as logistic regression, gamma set as 0, learning rate set as 0.2, maximum delta step set as 0, maximum depth set as 4, minimum sum of instance weight needed in a child set as 0, L2 regularization parameter lambda set as 7 and subsample set as 1.
5. <u>SVM:</u> Support Vector Machine was trained with ‘poly’ kernel, degree set as 3, gamma set as scale, regularization parameter C as 5 and maximum iterations set as 500,000.
6. <u>Decision Trees:</u> Were trained with criterion set as ‘entropy’, maximum depth set as 9, minimum number of samples required to be at a leaf node set as 9, minimum number of samples required to split an internal node set as 2 and splitter set as ‘random’.

### Testing

Stratified five-fold cross validation was used to acquire five models that were trained and validated on different folds. These models were then tested on the test set, and the results were averaged to determine model predictive performance.

#### Test set processing

The test set was first normalized using Min-Max scaling that was fit on the train set. Some of the rows had missing values, so we processed them in two ways as follows:

1. Rows missing values for any four or more of the five selected features were eliminated. The missing values of each sample in the resulting test set were imputed with the average of the values of its corresponding 10 nearest neighbours in the train set using KNN, where nearest neighbours were found with respect to the selected five features only. The nearest neighbours were determined through the inverse of euclidean distance between the data-points.
2. All the rows missing any one of the five values were dropped. This set has no imputation, and only includes the rows that have had all the values. This produces a test set which can simulate 100% real life testing scenario.

#### Testing on three cases

For realizing the true predictive performance and its consistency, the models were tested using three cases. Each of the following three cases have their own significance:

1. <u>Case ≤n:</u> If only *n* or less days are left till outcome Test samples that had the value of ‘Number of days till Outcome’ as greater than *n* ^*th*^ day were dropped. Then, testing was done on the cumulation of the rest of the samples.
2. <u>Case ≥n:</u> For *n* or more days in advance Test samples that had the value of ‘Number of days till Outcome’ as lesser than *n*^*th*^ day were dropped. Then, testing was done on the cumulation of the rest of the samples.
3. <u>Case =n:</u> On exactly *n* days before the outcome Test samples that had the value of ‘Number of days till Outcome’ equal to *n*^*th*^ day were chosen.

## Data Availability

All data is made available in the main manuscript and supplementary material.

## Acknowledgements

We thank Department of Science and Technology - Science and Enginering Research Board (grant no. CVD/2020/000343) for financial assistance.

