## Supplementary Material for "Machine Learning Based Clinical Decision Support System for Early COVID-19 Mortality Prediction"

.

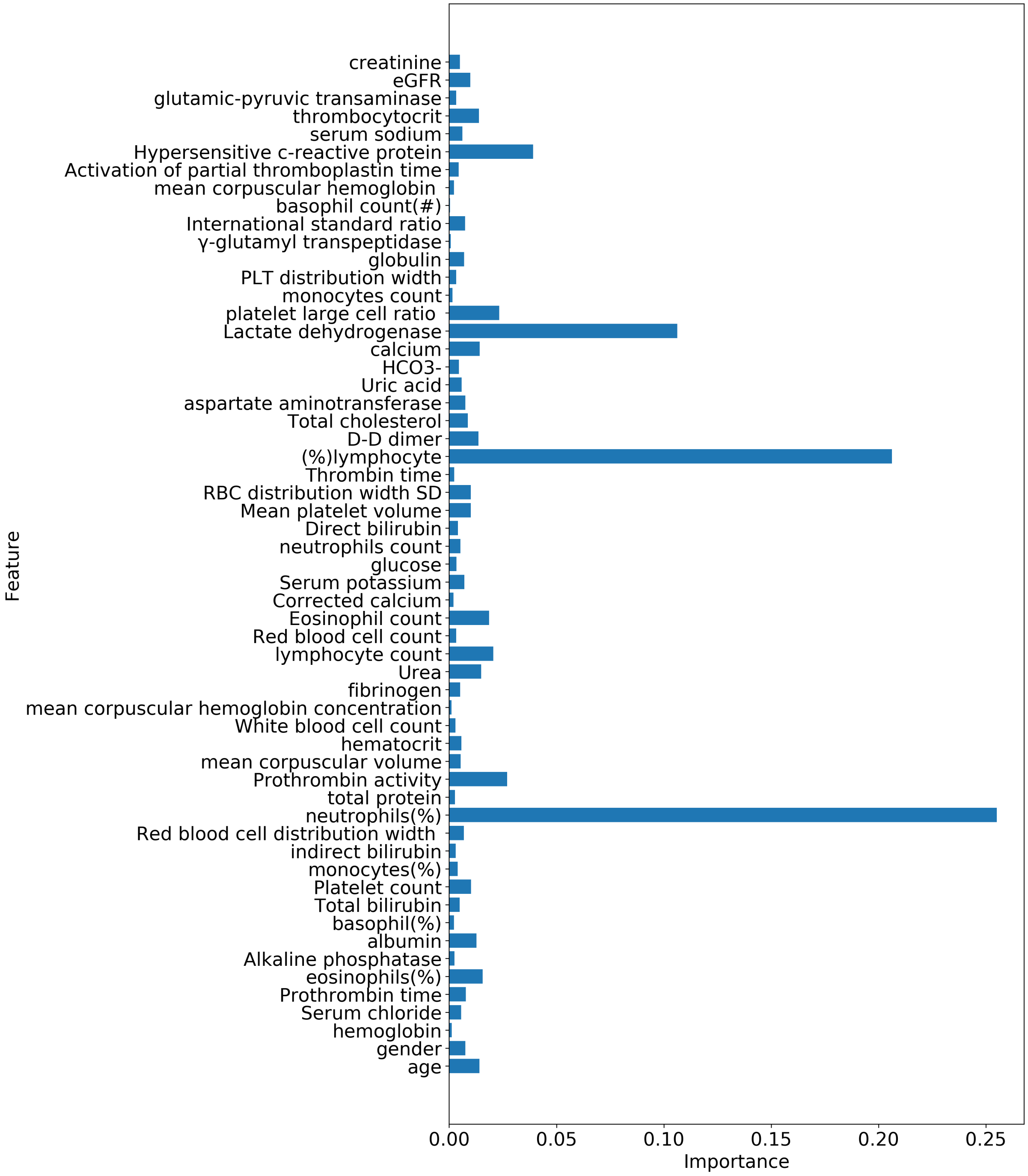


**Figure S1.** Mean relative importance of all the features in the train set determined using XGBoost. It is shown that the top four important features are neutrophils (%), lymphocyte (%), LDH and hs-CRP

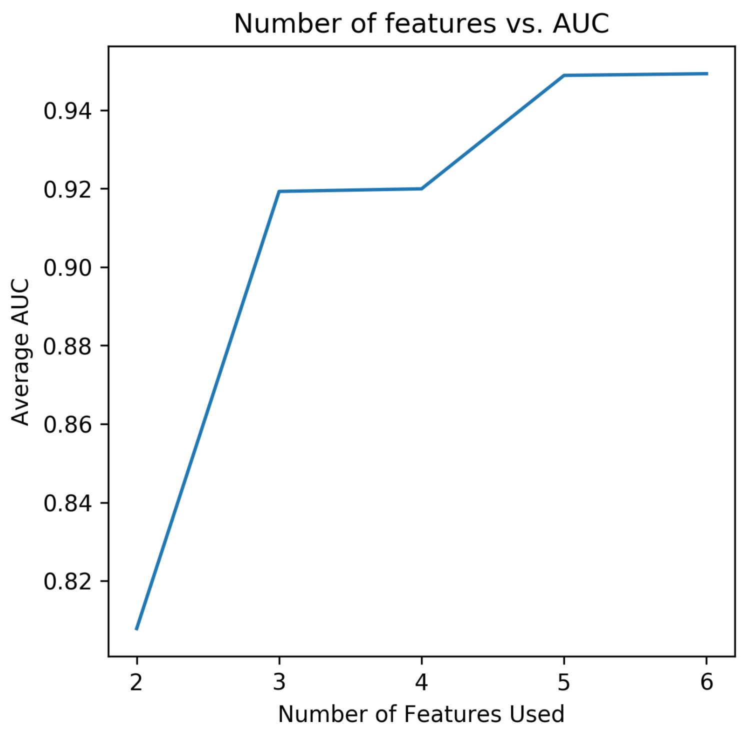


**Figure S2.** Number of features chosen vs. average AUC score obtained using neural network for feature selection. This suggests using the set of first five features after ordering the features in descending order of their relative importance. Selected five features are: age, neutrophils (%), lymphocytes (%), LDH and hs-CRP

| Model | Mean Accuracy% (std) | Mean F1 Score (std) | Mean AUC (std) |
|---|---|---|---|
| Neural Net | 96.526 (0.637) | 0.9687 (0.006) | 0.9895 (0.0057) |
| SVM | 95.305 (0.514) | 0.9577 (0.0046) | 0.9903 (0.0014) |
| Logistic Regression | 94.929 (0.188) | 0.9537 (0.0018) | 0.9934 (0.00015) |
| Random Forests | 94.178 (0.703) | 0.9467 (0.0065) | 0.9858 (0.0020) |
| XGBoost | 93.709 (1.3477) | 0.9435 (0.0118) | 0.9838 (0.0022) |
| Decision Tree | 90.892 (1.7963) | 0.914 (0.0182) | 0.9771 (0.0033) |

**Table S1.** Performance of various algorithms on the imputed test set

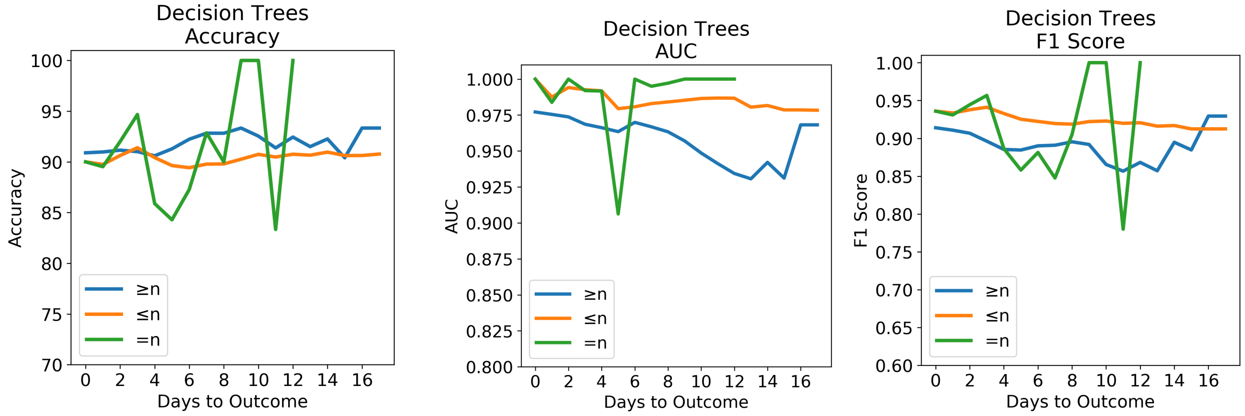


**(a)** Accuracy **(b)** AUC **(c)** F1-Score

**Figure S3.** The performance of Decision Trees on the imputed test set using the three testing cases. (Cases ≥n, ≤n and =n) (a) Accuracy of model evaluated for different days to outcome. (b) AUC score of model evaluated for different days to outcome. (c) F1 score of model evaluated for different days to outcome.

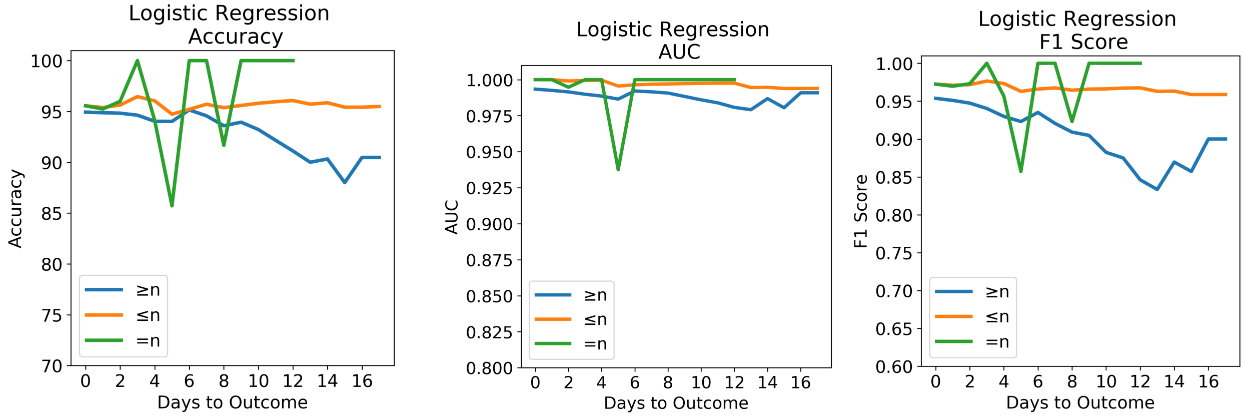


**(a)** Accuracy **(b)** AUC **(c)** F1-Score

**Figure S4.** The performance of Logistic Regression on the imputed test set using the three testing cases. (Cases ≥n, ≤n and =n) (a) Accuracy of model evaluated for different days to outcome. (b) AUC score of model evaluated for different days to outcome. (c) F1 score of model evaluated for different days to outcome.

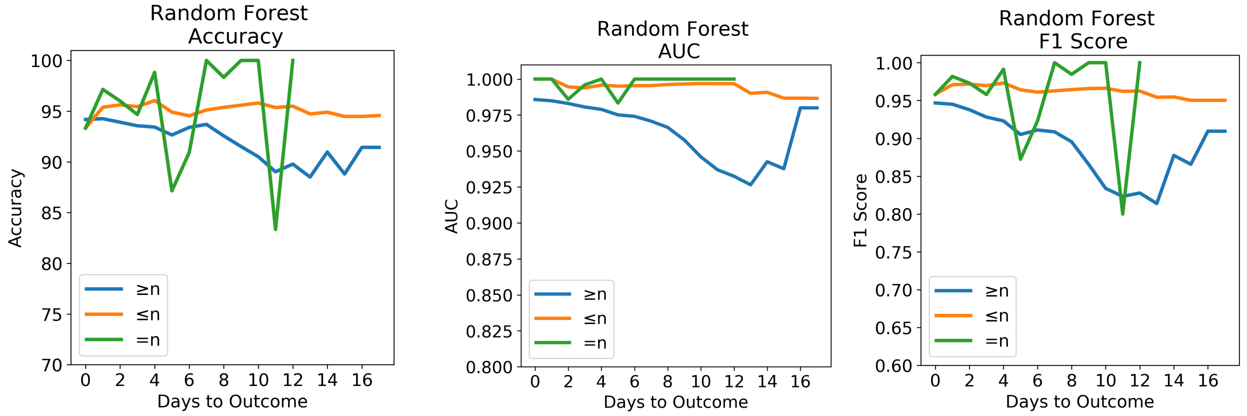


**(a)** Accuracy **(b)** AUC **(c)** F1-Score

**Figure S5.** The performance of Random Forests on the imputed test set using the three testing cases. (Cases ≥n, ≤n and =n) (a) Accuracy of model evaluated for different days to outcome. (b) AUC score of model evaluated for different days to outcome. (c) F1 score of model evaluated for different days to outcome.

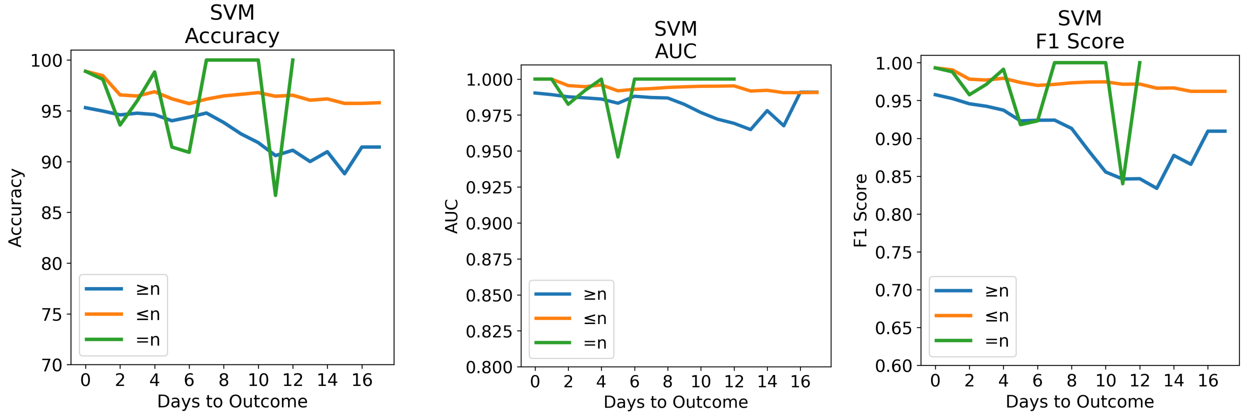


**(a)** Accuracy **(b)** AUC **(c)** F1-Score

**Figure S6.** The performance of SVM on the imputed test set using the three testing cases. (Cases ≥n, ≤n and =n) (a) Accuracy of model evaluated for different days to outcome. (b) AUC score of model evaluated for different days to outcome. (c) F1 score of model evaluated for different days to outcome.

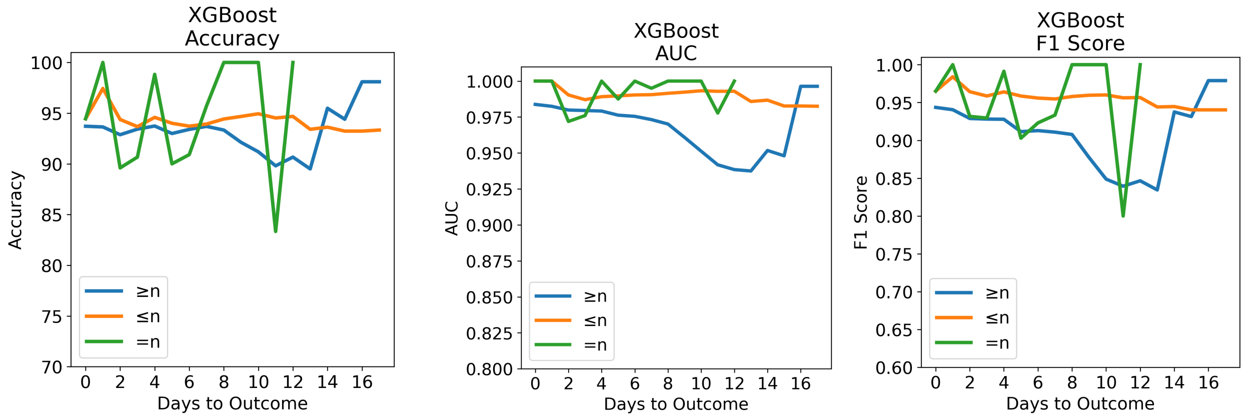


**(a)** Accuracy **(b)** AUC **(c)** F1-Score

**Figure S7.** The performance of XGBoost on the imputed test set using the three testing cases. (Cases $\geq$n, $\leq$n and =n) (a) Accuracy of model evaluated for different days to outcome. (b) AUC score of model evaluated for different days to outcome. (c) F1 score of model evaluated for different days to outcome.

| Model | Mean Accuracy% (std) | Mean F1 Score (std) | Mean AUC (std) |
|---|---|---|---|
| Neural Net | 94.608 (0.852) | 0.946 (0.009) | 0.982 (0.0086) |
| Logistic Regression | 93.043 («0.01) | 0.9298 («0.01) | 0.987 (0.00044) |
| SVM | 92.348 (0.852) | 0.9239 (0.0086) | 0.980 (0.0032) |
| Random Forests | 91.826 (1.301) | 0.917 (0.013) | 0.972 (0.004) |
| XGBoost | 90.608 (2.0132) | 0.9085 (0.0182) | 0.969 (0.0026) |
| Decision Tree | 89.0435 (2.101) | 0.8863 (0.0223) | 0.9615 (0.0057) |

**Table S2.** Performance of various algorithms on test set without imputation

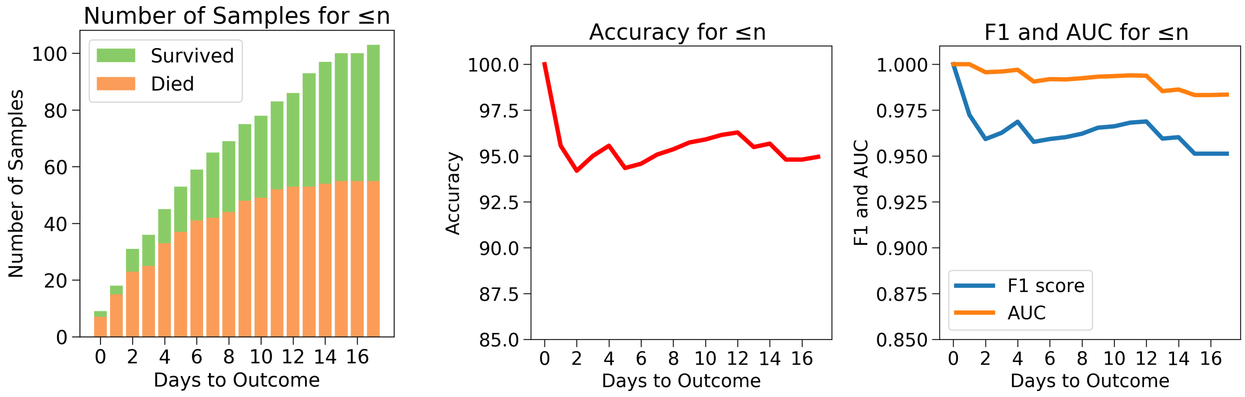


**(a)** Sample Distribution **(b)** Accuracy **(c)** F1-Score and AUC

**Figure S8.** Case 1: Number of days to outcome less than or equal to $n$. (a) The class-wise distribution of the cumulated data-points ($\leq n^{th} day$) for all samples in the test set without imputation. (b) Accuracy of the model evaluated for different days to outcome. (c) F1-score and AUC of the model evaluated for different days to outcome.

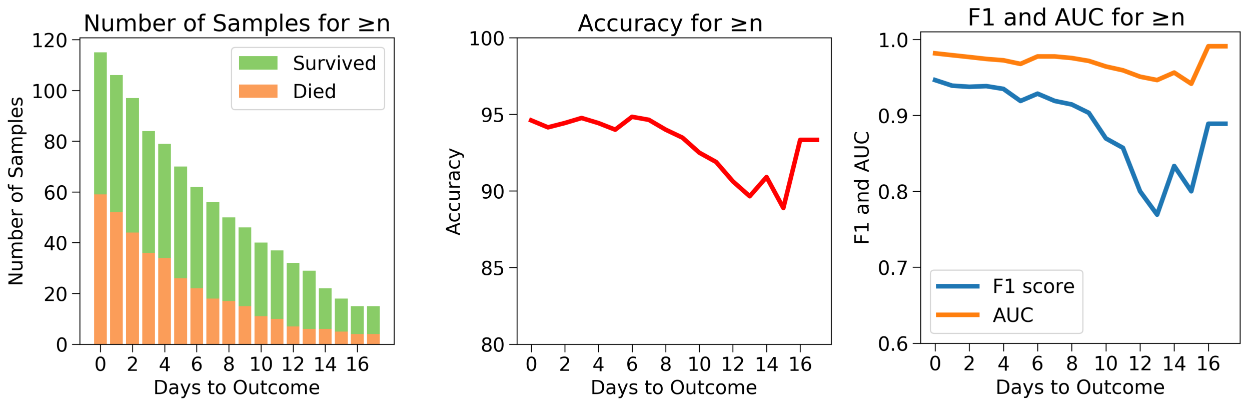


**(a)** Sample Distribution **(b)** Accuracy **(c)** F1-Score and AUC

**Figure S9.** Case 2: Number of days to outcome greater than or equal to $n$. (a) The class-wise distribution of the cumulated data-points ($\leq n^{th} day$) for all samples in the test set without imputation. (b) Accuracy of the model evaluated for different days to outcome. (c) F1-score and AUC of the model evaluated for different days to outcome.

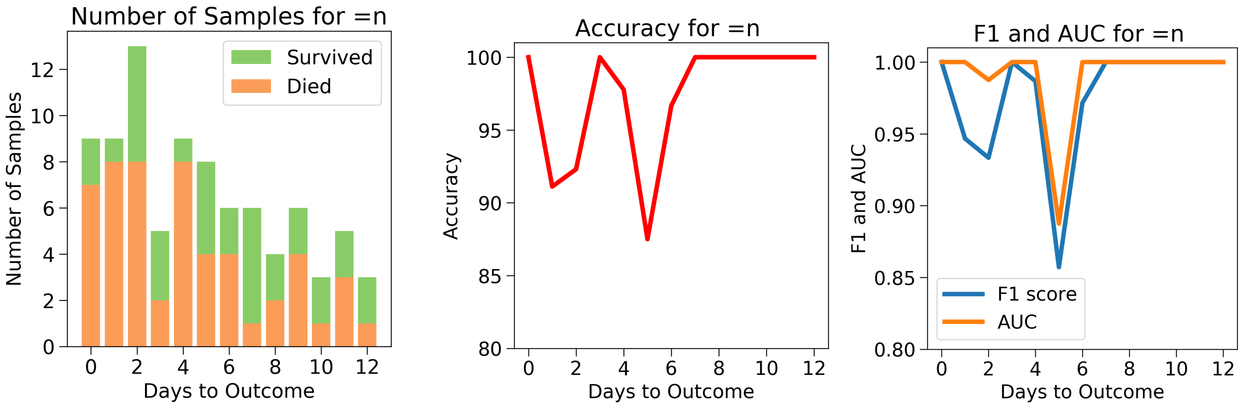


**(a)** Sample Distribution **(b)** Accuracy **(c)** F1-Score and AUC

**Figure S10.** Case 3: Number of days to outcome equal to $n$. (a) The class-wise distribution of the cumulated data-points ($\leq n^{th} day$) for all samples in the test set without imputation. (b) Accuracy of the model evaluated for different days to outcome. (c) F1-score and AUC of the model evaluated for different days to outcome.

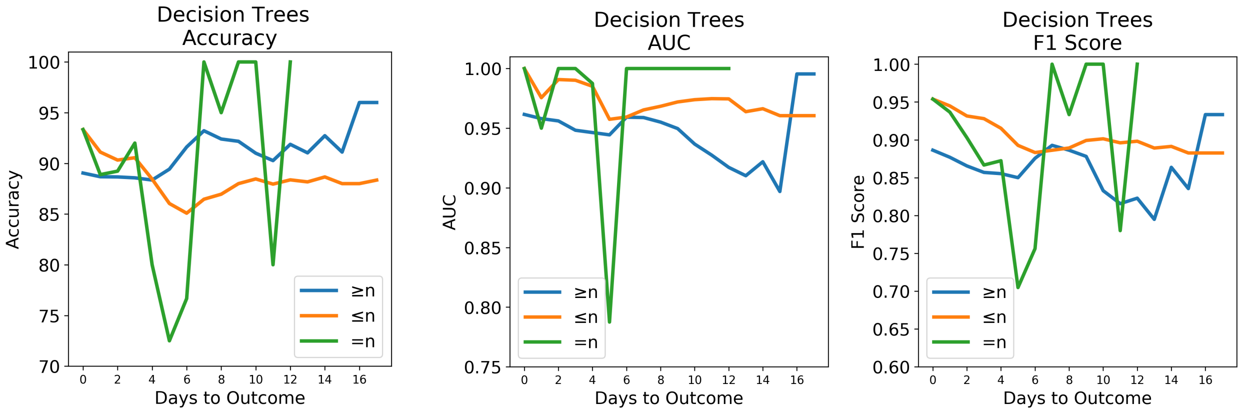


**(a)** Accuracy **(b)** AUC **(c)** F1-Score

**Figure S11.** The performance of Decision Trees on the test set without imputation using the three testing cases. (Cases $\geq$n, $\leq$n and =n) (a) Accuracy of model evaluated for different days to outcome. (b) AUC score of model evaluated for different days to outcome. (c) F1 score of model evaluated for different days to outcome.

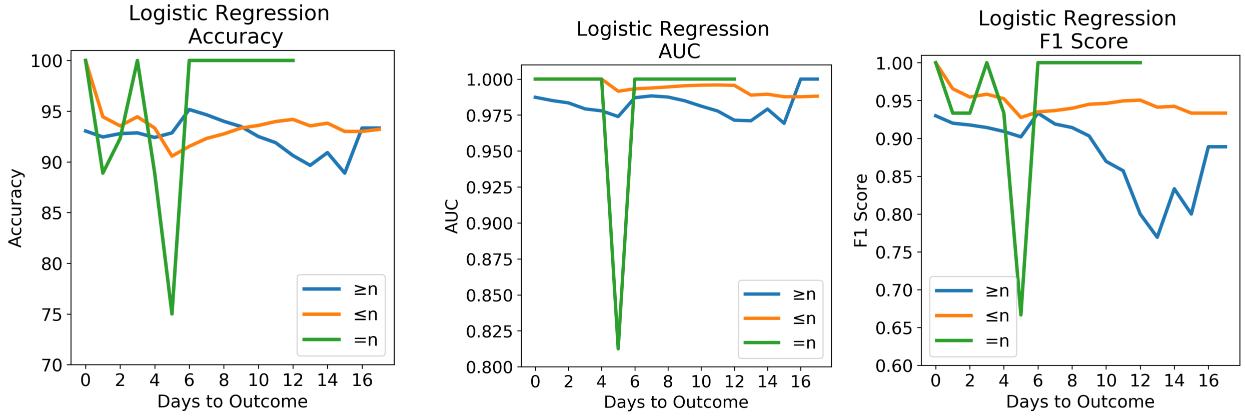


**(a)** Accuracy **(b)** AUC **(c)** F1-Score

**Figure S12.** The performance of Logistic Regression on the test set without imputation using the three testing cases. (Cases $\geq$n, $\leq$n and =n) (a) Accuracy of model evaluated for different days to outcome. (b) AUC score of model evaluated for different days to outcome. (c) F1 score of model evaluated for different days to outcome.

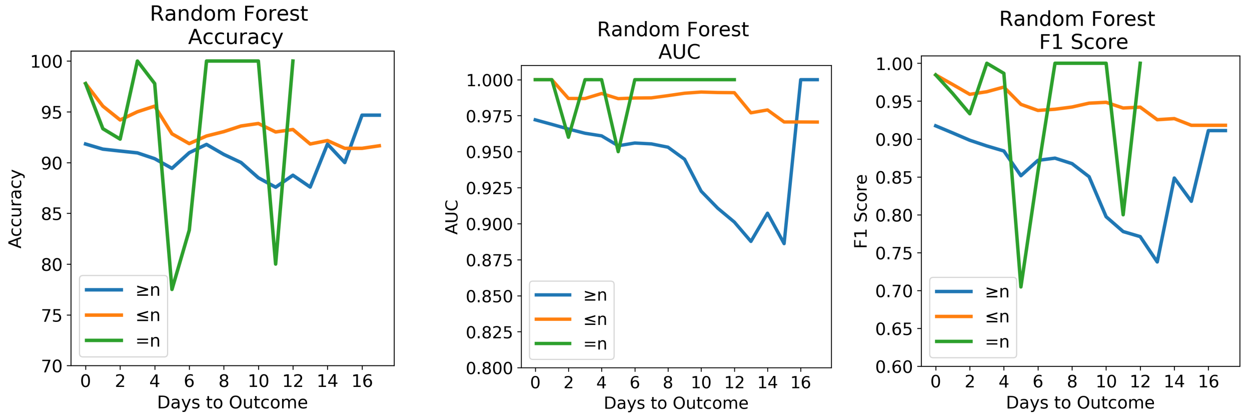


**(a)** Accuracy **(b)** AUC **(c)** F1-Score

**Figure S13.** The performance of Random Forests on the test set without imputation using the three testing cases. (Cases $\geq$n, $\leq$n and =n) (a) Accuracy of model evaluated for different days to outcome. (b) AUC score of model evaluated for different days to outcome. (c) F1 score of model evaluated for different days to outcome.

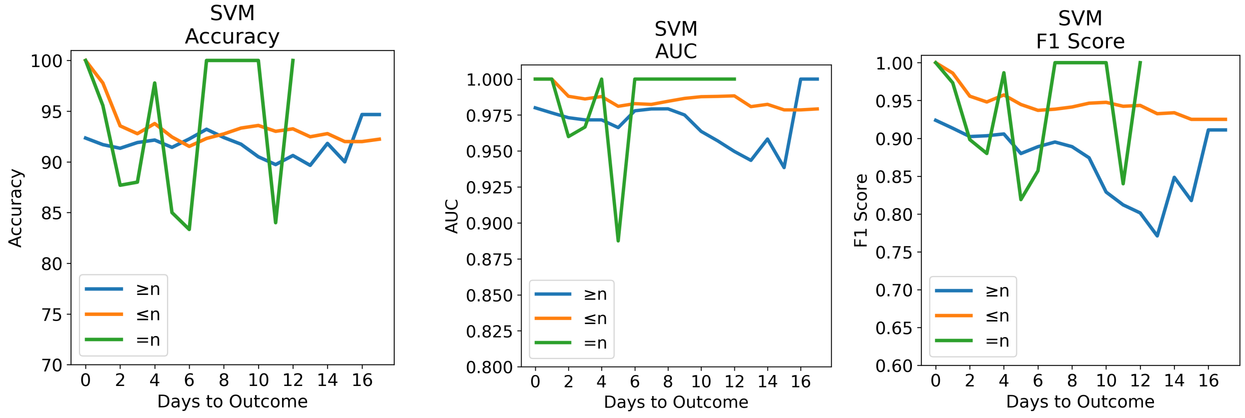


**(a)** Accuracy **(b)** AUC **(c)** F1-Score

**Figure S14.** The performance of SVM on the test set without imputation using the three testing cases. (Cases $\geq$n, $\leq$n and =n) (a) Accuracy of model evaluated for different days to outcome. (b) AUC score of model evaluated for different days to outcome. (c) F1 score of model evaluated for different days to outcome.

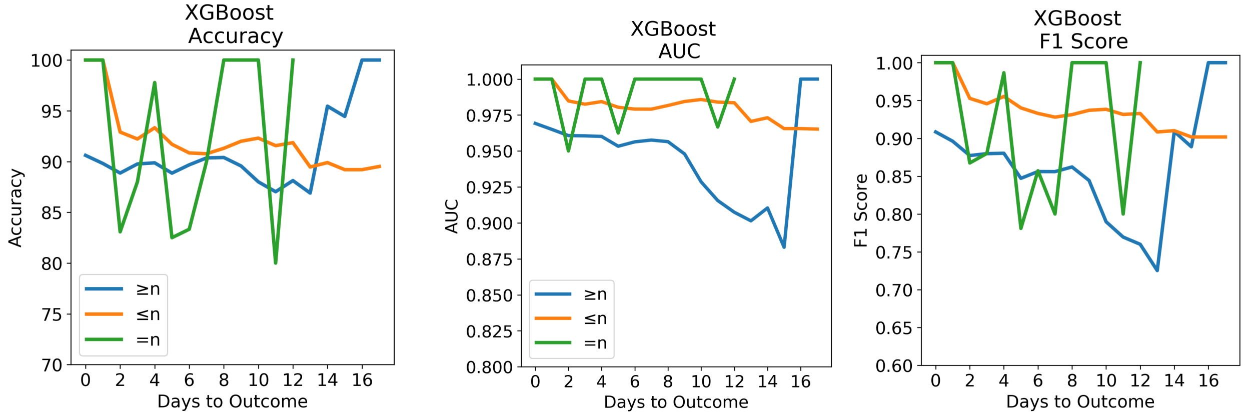


**(a)** Accuracy **(b)** AUC **(c)** F1-Score

**Figure S15.** The performance of XGBoost on the test set without imputation using the three testing cases. (Cases $\geq$n, $\leq$n and =n) (a) Accuracy of model evaluated for different days to outcome. (b) AUC score of model evaluated for different days to outcome. (c) F1 score of model evaluated for different days to outcome.

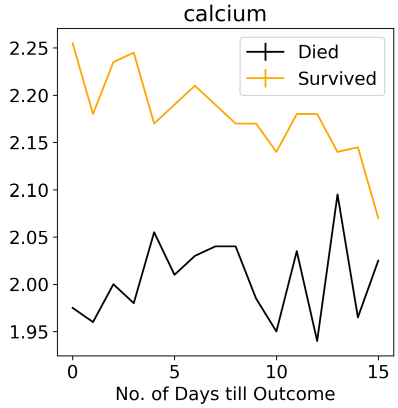


**(a)** Calcium

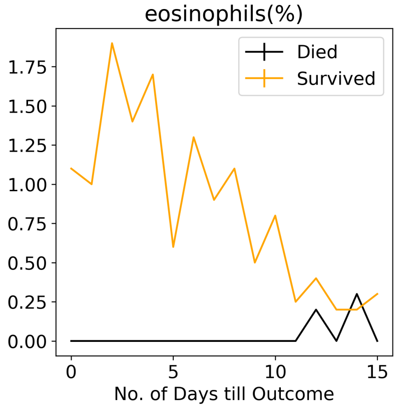


**(b)** Eosinphils (%)

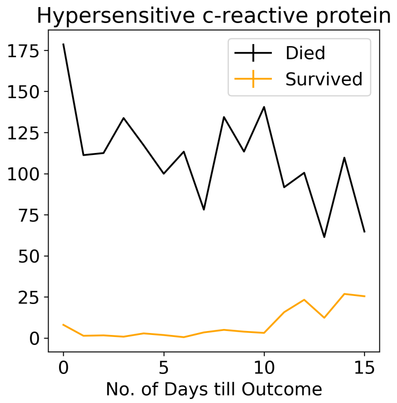


**(c)** Hs-CRP

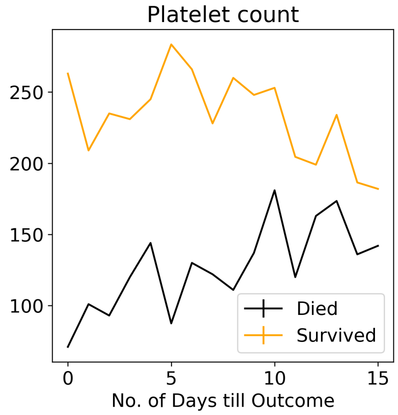


**(d)** Platelet Count

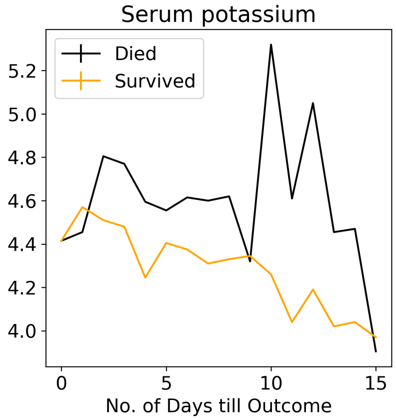


**(e)** Serum Potassium

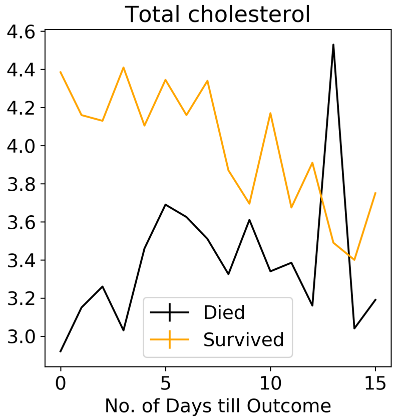


**(f)** Total Cholesterol

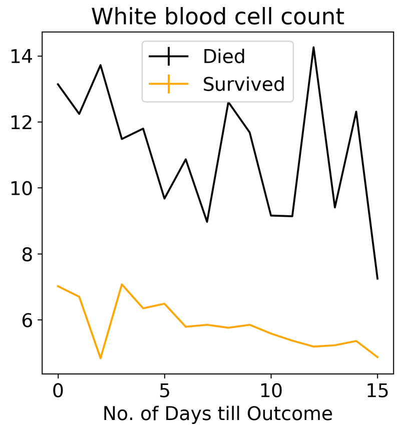


**(g)** WBC Count

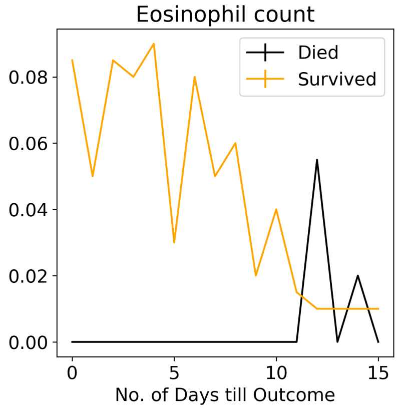


**(h)** Eosinophil Count

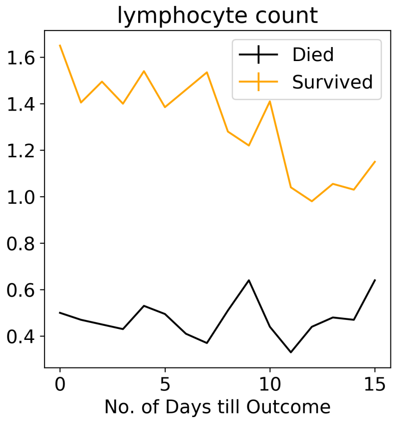


**(i)** Lymphocyte Count

**Figure S16.** Potential Features to predict number of days left to outcome: The median of features is determined on the set of data-points having the same number of days to outcome.

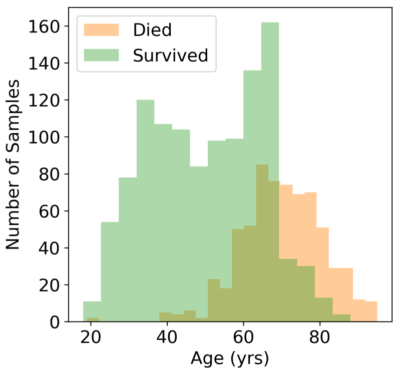


**(a)** Age

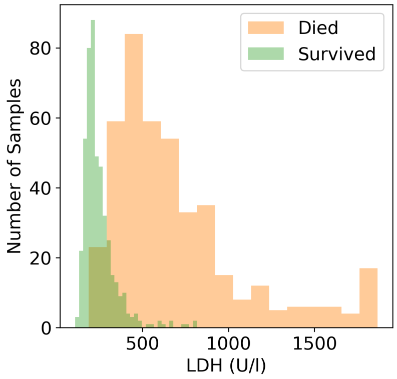


**(b)** LDH

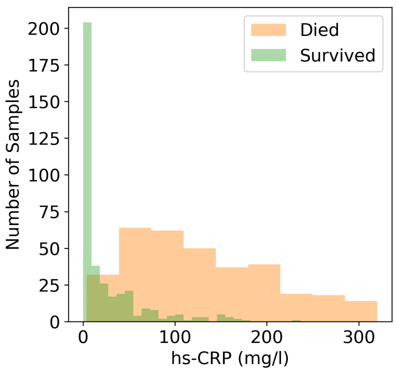


**(c)** hs-CRP

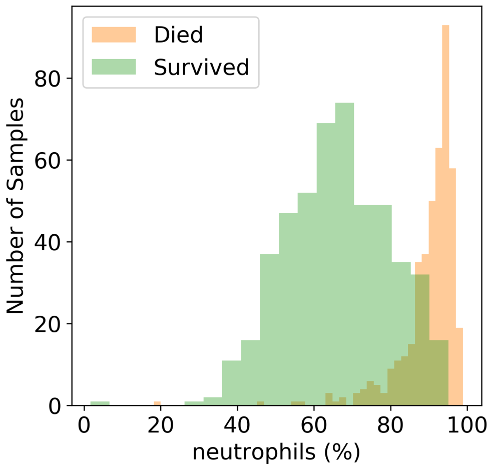


**(d)** Neutrophils (%)

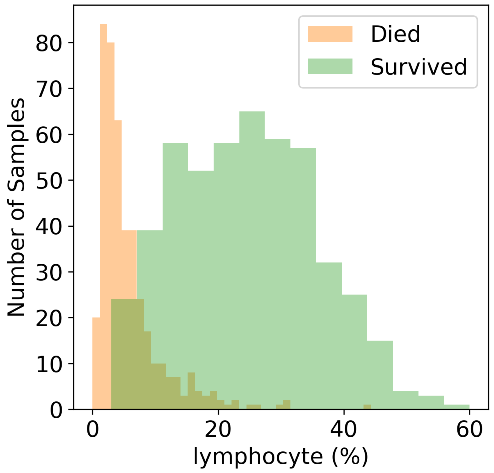


**(e)** Lymphocytes (%)

**Figure S17.** Distribution of the five selected features with respect to both the classes- survived and dead.
